# Genome-wide Meta-analysis of Alcohol Use Disorder in East Asians

**DOI:** 10.1101/2021.09.17.21263732

**Authors:** Hang Zhou, Rasmon Kalayasiri, Yan Sun, Yaira Z. Nuñez, Hong-Wen Deng, Xiang-Ding Chen, Amy C. Justice, Henry R. Kranzler, Suhua Chang, Lin Lu, Jie Shi, Kittipong Sanichwankul, Apiwat Mutirangura, Robert T. Malison, Joel Gelernter

## Abstract

**BACKGROUND:** Alcohol use disorder (AUD) is a leading cause of death and disability worldwide. Genome-wide association studies (GWAS) have identified ∼30 AUD risk genes in European populations, but many fewer in East Asians.

**METHODS:** We conducted GWAS and genome-wide meta-analysis of AUD in 13,551 subjects with East Asian ancestry, using published summary data and newly genotyped data from four cohorts: 1) electronic health record (EHR)-diagnosed AUD in the Million Veteran Program (MVP)sample; 2) DSM-IV diagnosed alcohol dependence (AD) in a Han Chinese-GSA (array) cohort;3) AD in a Han Chinese-Cyto (array) cohort; and 4) two AD datasets in a Thai cohort. The MVP and Thai samples included newly genotyped subjects from ongoing recruitment. In total, 2,254 cases and 11,297 controls were analyzed. An AUD polygenic risk score was analyzed in an independent sample with 4,464 East Asians (Kaiser Permanente data from dbGaP). Phenotypes from survey data and ICD-9-CM diagnoses were tested for association with the AUD PRS.

**RESULTS:** Two risk loci were detected: the well-known functional variant rs1229984 in *ADH1B* and rs3782886 in *BRAP* (near the *ALDH2* gene locus) are the lead variants. AUD PRS was significantly associated with days per week of alcohol consumption (beta = 0.43, se = 0.067, p = 2.47×10^−10^) and nominally associated with pack years of smoking (beta = 0.09, se = 0.05, p = 4.52×10^−2^) and ever vs. never smoking (beta = 0.06, se = 0.02, p = 1.14×10^−2^).

**CONCLUSIONS:** This is the largest GWAS of AUD in East Asians to date. Building on previous findings, we were able to analyze pleiotropy, but did not identify any new risk regions, underscoring the importance of recruiting additional East Asian subjects for alcohol GWAS.

## Introduction

Globally, alcohol use disorders (AUD) are among the top causes of morbidity and mortality [1]. Numerous factors predispose to the risk of developing AUD. Genetic factors contribute substantial risk to the etiology of AUD [2], and the heritability has been estimated to be ∼0.5 in twin studies [3]. Genome-wide association studies (GWAS) of AUD have been completed in multiple populations including European (EUR), African, Latin American and Asians [4-11]. To date, the largest GWAS of problematic alcohol use (PAU, a proxy for AUD) in 435,563 EUR subjects identified 29 independent risk variants [12]. In contrast, the largest GWAS of AUD in East Asians included less than 1% of this number: 3,381 subjects [13]. Genetic architecture often differs between populations; polygenic risk prediction between populations, though sometimes useful, often is not transferable [14, 15]. Thus, it is critically important that non-EUR populations be investigated to permit inferences to be made about these ancestral populations, which represent the majority of the world’s people [16, 17].

Because of the limited sample available and consequent lack of power for GWAS, little is known about the genetic architecture of AUD in East Asians. The most consistent loci identified are *ADH1B* (alcohol dehydrogenase 1b) and *ALDH2* (aldehyde dehydrogenase). Candidate studies of *ADH1B*\*rs1229984 and *ALDH2*\*rs671 in East Asians showed strong association between these functional variants and alcohol dependence [18, 19]. The first GWAS of alcohol dependence (AD) in a Chinese sample was conducted in 102 male cases and 212 male controls; rs3782886 in the *ALDH2* region was genome-wide significant (GWS) [5] despite the very small sample size. The first GWAS of AD in Thai samples included 1,045 subjects and identified rs149212747 in the *ALDH2* region as the lead variant [6]. The latest GWAS of AUD in a Chinese cohort identified both *ADH1B* and *ALDH2* genes as risk loci [13]. However, only a small proportion of the variance was explicable by variants in these genes. Larger samples are required to identify more risk variants to provide a better understanding of the genetic architecture in Asian populations.

Here we conducted a GWAS that combined 4 datasets from previously published cohorts and newly genotyped subjects from Thai and MVP cohorts. In total, 13,551 subjects of East Asian ancestry were analyzed, including 2,254 AUD cases. We then analyzed the resulting AUD PRS in an independent East Asian sample for associations with 26 phenotypes from surveys or ICD diagnoses. This GWAS of AUD is the largest to date in East Asians.

## Methods and Materials

### Datasets

#### Thai METH – GSA

As described previously [6], subjects were recruited in two stages for studies of the genetics of methamphetamine dependence (Thai METH). For both stages, subjects were recruited in Bangkok and assessed using the Thai version of the Semi-Structured Assessment for Drug Dependence and Alcoholism (SSADDA) [20]. The IRB protocols were approved by both the Chulalongkorn University (Thailand) IRB and the Yale University Human Research Protection Program. All subjects provided written informed consent prior to their research participation.

The first stage included methamphetamine users hospitalized between 2007 and 2011 for 4 months of residential drug treatment (Thai METH-GSA, Table 1) [21]. DNA samples were genotyped on the Illumina (San Diego, CA) Global Screening Array (GSA). Among the 863 genotyped subjects, we removed those with sample genotype call rate < 0.9, mismatched genotypic and phenotypic sex, or excess heterozygosity rate [6]. Unlike for our prior report, here we retained related subjects and applied linear mixed models to correct for relatedness (see below). SNPs with genotype call rate ≥ 0.95, minor allele frequency (MAF) ≥ 0.01, or Hardy-Weinberg Equilibrium (HWE) p-value > 10^−6^ were kept for imputation. Imputation was done by IMPUTE2 [22] with 1000 Genome project phase 3 data [23] as reference. SNPs with imputation INFO score ≥ 0.8 were retained for association analyses. Principal component analysis (PCA) was performed for the remaining subjects using EIGENSOFT [24, 25]. In contrast with our previous study, here we used DSM-IV alcohol dependence (AD) to define case status, rather than the DSM AUD criterion count to match the design in other cohorts. This yielded 127 cases and 405 were exposed controls. Linear mixed models implemented in GEMMA [26] were used to test association, with age, sex, and the first 10 PCs as covariates.

**Table 1.** Sample characteristics.

| **Cohorts** | **Traits** | **N (%female)** | **Mean (SD), Age** | **# Cases** |
| --- | --- | --- | --- | --- |
| Thai METH – GSA | DSM-IV AD | 532 (49.4) | 26.6 (6.9) | 127 |
| Thai METH – MEGA | DSM-IV AD | 2,370 (42.5) | 34.7 (10.1) | 794 |
| MVP – EAA | ICD 9/10 AUD | 6,955 (10.7) | 53.4 (17.1) | 701 |
| Han Chinese – GSA | DSM-IV AD | 3,381 (29.9) | 34.2 (8.3) | 533 |
| Han Chinese – Cyto | DSM-IV AD | 313 (0) | 49.6 (14.7) | 99 |
| Total |  | 13,551 |  | 2,254 |

#### Thai METH – MEGA

Second-stage subjects (N = 3,161; the Thai METH-MEGA sample, Table 1) were recruited from 2015 to 2020 [6]. DNA samples were genotyped using the Illumina Multi-Ethnic Global Array (MEGA). We removed subjects with sample genotype call rate < 0.95, sex mismatch, excess heterozygosity rate, or that were duplicates. SNPs with genotype call rate ≥ 0.95, or MAF ≥ 0.01, or HWE p-value > 10^−6^ were retained for imputation as with the Thai METH-GSA sample. We included 794 cases and 1576 alcohol-exposed controls in the association analysis, which used GEMMA and age, sex and the first 10 PCs as covariates.

#### MVP – EAA

The Million Veteran Program (MVP) is an ongoing observational cohort study and mega-biobank supported by the U.S. Department of Veterans Affairs (VA) [27, 28]. In October 2020, MVP released the latest genotype data (Release 4), which included 658,582 subjects. Quality control was first done by the MVP Release 4 Data Team and included the removal of duplicate DNA samples and those with sex mismatch, excessive heterozygosity, or a genotype call rate < 0.985. We ran PCA for the MVP subjects with 1000 Genome Project phase 3 (1KG) as the reference, defining genetic groups based on the top 10 PCs [8]. For subjects grouped as East Asian Americans (EAA), we ran a second PCA and removed outliers with PC scores >6 standard deviations from the mean on any of the 10 PCs, yielding in 7,364 EAAs. Imputation [22] was performed specifically for the EAAs using the 1KG as reference. SNPs with genotype call rate ≥ 0.95, MAF ≥ 0.01, HWE > 1×10^−6^, and imputation INFO ≥ 0.8 were retained for analysis. As for our prior study in EUR [12], subjects with ≥ 2 outpatient or ≥ 1 inpatient International Classification of Diseases (ICD) codes for AUD were defined as cases (N = 701, Table 1) and subjects with no AUD ICD code as controls (N = 6,254). BOLT-LMM [29] was used to correct for relatedness, with age, sex, and the first 10 PCs as covariates.

#### Han Chinese – Cyto

This first GWAS of alcohol dependence, flushing response, and maximum daily drinks consumed in a Han Chinese family sample [5] used the Illumine Cyto12 array (Table 1). Whereas the cohort was not imputed in the original report, we re-analyzed the data and imputed the SNPs for an AD GWAS. Subjects with genotype call rate < 0.95, duplicated DNA samples, mismatched sex, or excessive heterozygosity were removed, resulting in 511 subjects for imputation. Imputation used IMPUTE2 and 1KG reference, SNPs with minor allele frequency < 0.01, genotype call rate < 0.95, HWE p-value < 1×10^−6^, or imputation INFO <0.8 were removed from further analyses. Due to the drinking practices and characteristic of this particular population that result in few AD cases in females, only males were included in this analysis. After QC, 99 DSM-IV-diagnosed male AD cases and 214 male alcohol-exposed controls were analyzed using GEMMA to correct for relatedness, with age, sex and 10 PCs as covariates.

#### Han Chinese – GSA

The second case-control AD GWAS in Han Chinese [13] included 533 cases and 2,848 alcohol-exposed controls who were genotyped using the GSA array (Table 1). Here we used the summary statistics from this study.

### Meta-analysis

Using association analyses or summary statistics for each of the four cohorts, effective sample-size weighted meta-analysis was performed using METAL [30]. SNPs present in only one cohort or in less than 15% of the total samples were removed (6.8 million SNPs remained). To define lead variants, the meta-analysis summary data were clumped by LD with r^2^ < 0.1 in a 2500-kb window, using 1KG East Asians as the LD reference. Additional conditional analyses [31] were performed to define independent variants when more than one lead variant was present in the same locus (e.g., the chr4 ADH gene region).

### Polygenic Risk Scores

#### Target dataset

We requested and downloaded from dbGaP (phs000788.v2.p3) data from the Kaiser Permanente Research Program on Genes, Environment, and Health (RPGEH) Genetic Epidemiology Research on Adult Health and Aging (GERA) cohort. This large and ethnically diverse cohort genotype data from 5,182 self-reported Asians using a custom Affymetrix Axiom array [32]. All subjects completed a broad written consent.

#### Imputation

Subjects with mismatched sex or genotype call rate < 0.95 were removed. The genomic build was transferred from 36 to 37 using LiftOver [33]. As we did for MVP, we ran PCA for the 5,182 Asian subjects using the 1KG as reference, clustering them into different groups. A second PCA among Asians was used to remove outliers, resulting in 4,464 genetically classified East Asians for imputation. Imputation was performed using IMPUTE2 and 1KG reference, SNPs with minor allele frequency < 0.01, genotype call rate < 0.95, HWE p-value < 1×10^−6^, or imputation INFO < 0.8 were removed from further analysis.

#### Target phenotypes

Two sources of phenotypes are included in this study. The first is survey data on physical observations, lifestyle and environment, including phenotypes such as BMI, general health, physical activity, alcohol use, smoking status and pack years. The second is ICD-9-CM. disease and conditions measures. Participant were coded as cases if there were at least two diagnoses in a disease category. Binary phenotypes with less than 100 cases were removed from analyses. See Table 2 for details of the target phenotypes.

**Table 2.** Tested phenotypes in GERA and association results with AUD PRS. If not specified for distribution, 1 is case and 0 is control.

| Traits | Distribution | Beta (se) | p-value |
| --- | --- | --- | --- |
| **Alcohol use in days per week \*\*** | 1=2757, 2=603, 3=503, 4=159, 5=267<sup>c</sup> | 0.43 (0.07) | **2.47×10<sup>-10</sup>** |
| **Smoking in pack years \*** | 0=3232, 1=530, 2=306, 3=128, 4=44<sup>d</sup> | 0.09 (0.05) | **4.52×10<sup>-2</sup>** |
| **Ever vs. never smoked \*** | 1=1055, 0=3232 | 0.06 (0.02) | **1.14×10<sup>-2</sup>** |
| Former vs. current smoker | 1=924, 0=131 | -0.04 (0.04) | 2.50×10<sup>-1</sup> |
| Physical activity | 1=897, 2=958, 3=1181, 4=1323<sup>e</sup> | 0.02 (0.06) | 7.97×10<sup>-1</sup> |
| Health status | 1=740, 2=1506, 3=1625, 4=476<sup>f</sup> | 0.03 (0.05) | 5.23×10<sup>-1</sup> |
| *Disease or conditions* |  |  |  |
| Acute reaction to stress | 1=275, 0=4189 | -0.01 (0.01) | 5.22×10<sup>-1</sup> |
| Allergic rhinitis | 1=1307, 0=3157 | 0.01 (0.03) | 5.62×10<sup>-1</sup> |
| Asthma | 1=654, 0=3810 | -0.01 (0.02) | 7.21×10<sup>-1</sup> |
| Cancer: any<sup>a</sup> | 1=529, 0=3935 | -0.00 (0.02) | 9.36×10<sup>-1</sup> |
| Cardiovascular disease: any<sup>b</sup> | 1=688, 0=3776 | -0.03 (0.02) | 1.48×10<sup>-1</sup> |
| Major depressive disorder | 1=262, 0=4202 | 0.01 (0.01) | 3.66×10<sup>-1</sup> |
| Dermatophytosis | 1=374, 0=4090 | -0.01 (0.02) | 4.87×10<sup>-1</sup> |
| Type II diabetes | 1=729, 0=3735 | 0.03 (0.02) | 9.65×10<sup>-2</sup> |
| Dyslipidaemia | 1=2192, 0=2272 | -0.02 (0.03) | 5.17×10<sup>-1</sup> |
| Hemorrhoids | 1=716, 0=3748 | 0.01 (0.02) | 6.19×10<sup>-1</sup> |
| Hernia abdominopelvic cavity | 1=177, 0=4287 | 0.00 (0.01) | 7.54×10<sup>-1</sup> |
| Hypertensive disease | 1=2028, 0=2436 | -0.00 (0.02) | 9.52×10<sup>-1</sup> |
| Insomnia | 1=185, 0=4279 | -0.01 (0.01) | 2.30×10<sup>-1</sup> |
| Iron deficiency anemias | 1=118, 0=4346 | 0.00 (0.01) | 9.14×10<sup>-1</sup> |
| Irritable bowel syndrome | 1=103, 0=4361 | 0.00 (0.01) | 7.05×10<sup>-1</sup> |
| Macular degeneration | 1=130, 0=4334 | -0.00 (0.01) | 7.94×10<sup>-1</sup> |
| Osteoarthritis | 1=941, 0=3523 | -0.01 (0.02) | 6.72×10<sup>-1</sup> |
| Osteoporosis | 1=392, 0=4072 | 0.01 (0.01) | 6.46×10<sup>-1</sup> |
| Psychiatric disorder: any | 1=433, 0=4031 | -0.00 (0.02) | 9.93×10<sup>-1</sup> |
| Peripheral vascular disease | 1=160, 0=4304 | 0.01 (0.01) | 3.81×10<sup>-1</sup> |

Note: <sup>a</sup>Cancer: includes malignant tumors, neoplasms, lymphoma and sarcoma. <sup>b</sup>Heart disease: includes ischemic heart disease, cardiac arrest, congestive heart failure, dysrhythmias, cardiomyopathy, aortic aneurysm, and cerebrovascular disease, but excludes PVD which is encompassed by the PVD variable. <sup>c</sup>Days of alcohol intake per week, 1 is no days, 2 is 1 day, 3 is 2-4 days, 4 is 5-6 days, 5 is every day. <sup>d</sup>Pack-years for former or current smoker, 0 = 0, 1 < 10, 2 = 10-20, 3 = 20-30, 4 ≥ 30. <sup>e</sup>Physical activity total metabolic equivalency of task (MET), 1 = first quartile, 0-173 for males and 0-74 for females, 2 = second quartile, 174-600 for males and 75-344 for females, 3 = third quartile, 601-1380 for males and 345-983 for females, 4 = fourth quartile, 1381+ for males and 984+ for females. <sup>f</sup>Health status, 1 = excellent, 2 = very good, 3 = good, 4 = fair.

#### Polygenic risk scoring and association

PRS-CS [34] was used to infer posterior effect sizes of SNPs using GWAS summary statistics for AUD from this study, and an external East Asian LD reference panel (generated by the authors of PRS-CS using the 1KG phase3 East Asian reference). We used PLINK v1.9 [35] for polygenic risk scoring in the target East Asian samples. GEMMA was used analyze associations between the PRS and target phenotypes, accounting for relatedness and correcting for age (in 5-year categories), sex and the first 10 PCs.

### Additional downstream analyses

We used LD score regression [36] to estimate the SNP-based heritability of AUD using 1KG East Asians as the LD reference. We also investigated the trans-ancestry genetic correlation between this study sample and PAU in EUR populations using popcorn, a method that uses only summary-level data from GWAS while accounting for LD [37]. Trans-population meta-analysis between this study and PAU in EUR was conducted using METAL. Multi-trait analysis [38] was performed, which combined data from this study with excessive alcohol consumption defined as weekly intake > 150 mL of alcohol for ≥ 6 months from the Taiwan Biobank [39]

## Results

### Genome-wide association and meta-analyses

As in our previous study of AUD in East Asians [13], in a meta-analysis here of 2,254 cases and 11,297 controls, we confirmed two loci that were significantly associated with AUD (Table 1, Figure 1). One locus is on chromosome 4q23 and includes multiple alcohol dehydrogenase genes. After LD clumping, there are two lead SNPs in this locus. The first is rs1229984 (Arg48His, p = 3.35×10^−17^) in *ADH1B* (Alcohol Dehydrogenase 1B (Class I), Beta Polypeptide), the second is rs1814125 (p = 2.14×10^−10^) near *ADH1C*. Conditional analysis indicated that rs1814125 is not independent from rs1229984. Another locus is a long region with high LD on chromosome 12 for which there is positive selection in East Asians, which covers *ALDH2* (Aldehyde Dehydrogenase 2) and *BRAP* (BRCA1 associated protein) genes. The lead SNP is rs3782886 (p = 1.68×10^−29^), a coding variant in the *BRAP* gene. No other associations were detected in this study.

**Figure 1.**
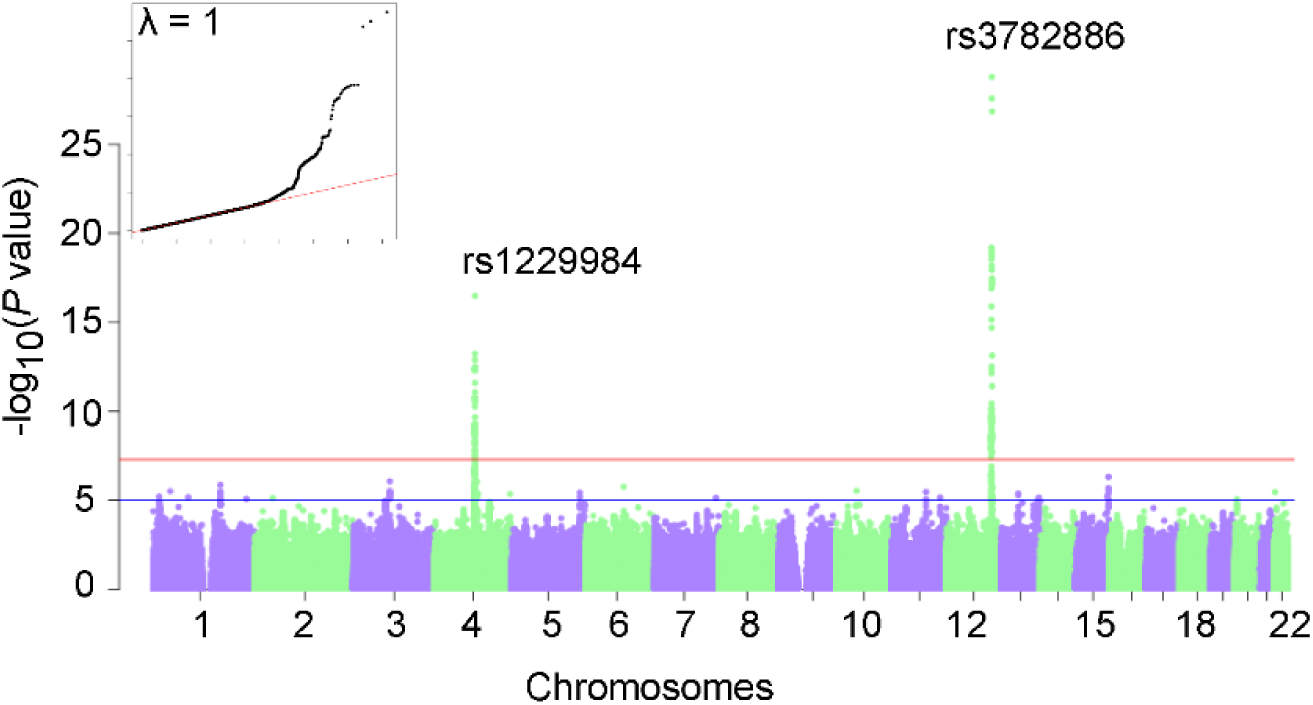
Manhattan plot of the meta-analysis of AUD in 13,551 East Asians.

### Polygenic risk score for AUD

We calculated PRS for AUD in an independent East Asian cohort from the GERA cohort. We tested 26 phenotypes from survey and ICD-9-CM diagnosed conditions for association with the AUD PRS (Table 2). As expected, AUD PRS is significantly associated with alcohol consumption as measured in days per week of drinking (beta = 0.43, se = 0.067, p = 2.47×10^−10^). Also, AUD PRS is nominally significantly associated with pack years of smoking (beta = 0.09, se = 0.05, p = 4.52×10^−2^) and ever vs. never smoking (beta = 0.06, se = 0.02, p = 1.14×10^−2^), but these associations did not survive multiple testing correction. None of the other traits in this small target sample were associated with AUD PRS. None of the additional downstream analyses provided significant results nor did they identify additional association signals.

## Discussion

We collected data from 13,551 subjects with East Asian ancestry to conduct the largest meta-analysis to date for an alcohol-related trait in this population (quadruple the previous largest reported sample). We detected association signals at the *ADH1B* and *ALDH2* loci with substantially stronger statistical significance than has been seen previously, but did not identify any novel risk loci. This is mostly consistent with observations from other studies, where GWAS of alcohol-related traits with sample sizes in this range are generally underpowered to detect multiple replicable variants [4, 7, 40-43]. In EUR and African-Ancestry populations, the first and strongest associations detected have been at *ADH1B*. The *ALDH2* association is, to this date, unique to Asians. It is a common issue for complex traits like AUD that many variants contribute to the heritability, each with a small effect size [44, 45]. Missing ancestral diversity in human genetic studies is a critical issue and recruitment of non-EUR subjects is crucial to addressing this disparity [16, 17]. The identification of *ALDH2*, which as noted is unique to Asians, exemplifies that there are differences in the genetic architecture of AUD between Asians and, for example, EUR, making well-powered investigations in this population an important scientific issue. Beyond identifying *ADH1B* and *ALDH2* with greater statistical significance than previous studies, the present investigation extends prior findings in several ways, including by examining the utility of the AUD PRS derived from this meta-analysis in an independent cohort of 4,464 East Asians and testing the association between the AUD PRS and alcohol, smoking, and other traits.

The two genes implicated – *ADH1B* and *ALDH2* – are involved in ethanol metabolism [45]. *ADH1B* encodes an alcohol dehydrogenase that oxidizes alcohol to acetaldehyde, which is then oxidized to acetate by aldehyde dehydrogenases, including that encoded by *ALDH2*. This is the major metabolic pathway for ethanol metabolism but other genes are involved as well. For example, in the first step, *ADH1C, ADH4* and *ADH7*, which map to the same chromosome 4 gene cluster as *ADH1B*, encode proteins that perform similar biological functions under certain conditions, *ALDH1A1* and *ALDH1B1* similarly encode proteins with roles that are sometimes overlapping with that of *ALDH2* [46]. Given the importance of other genes in the metabolic pathway, lead variants in genes other than *ADH1B*\*rs1229984 (EUR and Asian) and *ALDH2\**rs671 (Asian) have been reported [6, 8, 12, 13]. Some of these associations are supported by conditional analyses [8, 12], and some appear to be variants that “hitchhike” with rs1229984 or rs671 due to their strong LD. Here, conditional analyses identified only one lead variant at each locus: rs1229984 (p = 3.35×10^−17^) in the *AHD1B* region and rs3782886 (p = 1.68×10^−29^) in the *ALDH2* region. The high LD between rs3782886 and rs671 (r^2^ = 0.98) makes it difficult to distinguish the real causal variant, though biochemical analysis favors rs671 (reviewed in [45]), which is nearly a null variant.

Rs3782886 in the *BRAP* gene (breast cancer suppressor protein (BRCA1)-associated protein) has been associated with many traits in East Asians, include alcohol-related traits [39, 47], myocardial infarction [48], and a biochemical trait – alanine aminotransferase level [49]. Some or all these associations could be due to the high LD with rs671 (as in this study), or reflect effects on activity of the metabolic pathway or cerebral cortical neurogenesis (argued in [39]). We would suggest that the different lead variants (rs671 or rs3782886) in this high LD region could reflect uncertainty introduced by different SNP arrays, imputation processes, association analyses, or random variation in comparatively small samples. More data are needed to ascertain the true causal variant despite the previous support and mechanistic appeal of rs671.

We used additional analyses to explore the genetic architecture of AUD in East Asians. The SNP-based heritability estimate was very low with a large standard error (SE), indicating a lack of statistical power. Trans-ancestry genetic correlation between this study and PAU in EUR populations is 0.61 (se = 0.23, p = 0.10). The trans-population meta-analysis in which PAU in EUR was added did not detect any novel signals. Multi-trait analysis combining this study sample and excessive alcohol consumption from the Taiwan Biobank also identified no novel variants. Thus, additional study samples of European ancestry are needed to provide adequate power for GWAS of AUD in East Asians.

Since it is a genetically complex trait, we expect that there are many variants that contribute to the genetic risk of AUD, consistent with findings in EUR [12]. Polygenic risk score analysis is a powerful tool for the application of GWAS results to investigate associations with traits of interest, which has been used widely in studies to test the association with AUD or related phenotypes in target cohorts [7, 8, 12, 13]. Here, we analyzed AUD PRS from our meta-analysis in an independent East Asian cohort from GERA, a US cohort collected to facilitate research on the genetic and environmental factors that affect health and disease [32]. We tested the association between AUD PRS and 26 phenotypes in 4,464 subjects of East Asian ancestry. AUD PRS was significantly associated with alcohol consumption as measured using days of drinking per week (see Table 2), and nominally significantly associated with pack years of smoking and ever vs. never smoking, consistent with the shared genetic architecture of AUD and alcohol and (possibly) smoking traits in East Asians. These same, or closely similar, associations, have been well established in EUR [8, 12]. These was no association detected between AUD PRS and other diseases or conditions in this study.

This study has limitations, the most important of which is the sample size, which despite being the largest reported so far for East Asian provides limited statistical power. Second, the phenotypes among the different study samples are not identical, with AUD diagnosed as ICD-9/10 codes in MVP and DSM-IV AD in other cohorts. This analytic approach is supported by the high genetic correlation between AUD and AD in EUR, which is estimated to approach 1.0 [12]. Third, some cohorts used alcohol-exposed controls, and others used unscreened controls (i.e., the MVP). Controls with demonstrated exposure to alcohol are ideal, but such exposure is commonplace in all the populations studied. Finally, although all of the cohorts are of East Asian ancestry, there are population differences among cohorts that increase heterogeneity and reduce power. These include cultural or environmental differences that affect trait prevalence (e.g., drinking practices), and geographical differences that introduce genetic differences.

In conclusion, we conducted a GWAS of AUD in 13,551 East Asian subjects, in which we confirmed the two previously known risk loci and applied the AUD PRS in an independent cohort. Despite a large increment in sample size over the previous largest Asian-population GWAS, the power remains an important limitation. Accordingly, we will continue to recruit more East Asian subjects for alcohol studies and urge other investigators to do the same.

## Data Availability

The summary statistics can be shared upon request.

## Acknowledgements and Disclosures

This research used data from the Million Veteran Program, which was supported by funding from the Department of Veterans Affairs Office of Research and Development, Million Veteran Program Grant #I01BX003341. This publication does not represent the views of the Department of Veterans Affairs or the United States Government. Supported also by NIH (NIDA) R01 DA037974 (JG), and a NARSAD Young Investigator Grant from the Brain & Behavior Research Foundation (HZ). Data from the Kaiser Permanente Research Program on Genes, Environment, and Health (RPGEH) were accessed from dbGaP (phs000788.v2.p3).

